# The Health Impact of Electronic Nicotine Delivery Systems: A Systematic Review

**DOI:** 10.1101/2020.10.07.20208355

**Authors:** C Hajat, E Stein, S Shantikumar, R Niaura, P Ferrara, R Polosa

## Abstract

**Introduction:** The objective of this systematic review was to identify, report and critically appraise studies that have reported health outcomes from use of electronic nicotine delivery systems (ENDS).

**Methods:** We conducted a systematic review of all published literature on the health impact of ENDS products from 1^st^ January 2015 until February 1, 2020, following the PRISMA protocol, including across the databases, PubMed, Embase, Scopus and Google Scholar using medical subject headings.

A category for the level of evidence was assigned blindly using the Centres for Evidence Based Medicine framework. A similar approach was adopted to evaluate methodological quality of each study utilizing the National Institutes for Health (NIH) Quality Assessment Tools.

**Results:** The database search identified 755 studies and a further 265 were identified from other sources and reference reviews of which 37 studies met the eligibility criteria.

The majority of studies were of low strength for levels of evidence including 24 (65%) cross-sectional, 1(2.7%) case-control and six (16%) case studies. There were four (11%) cohort studies and only one (2.7%) RCT. There was only one (2.7%) meta-analysis or pooled study of observational study designs; there were no pooled results of randomized controlled trials. Of 37 studies, eight (22%) studies reported on benefits, two (2%) studies were neutral, reporting on both harm and benefits, the remaining 27 (73%) reported only on harms. The quality ratings were poor (20, 54%), fair (9, 24%) and good (8, 22%).

In our review ENDS use has not been shown to be causative for any CVD outcomes and has been shown to be beneficial for hypertensive patients. Switching from cigarettes to e-cigarettes resulted in reduced exacerbations of COPD, with no evidence of long-term deterioration in lung function. There was a suggestion of short-term reductions in respiratory function in asthmatics, but no increased risk of asthma in ENDS users who were never smokers. Mental Health, cancer and mortality have not been adequately studied to form any consensus with regards to health outcomes from ENDS use.

**Conclusion:** Our review suggests that the majority of studies on the use of ENDS products reported on negative health impacts with few reporting on health outcomes from switching from cigarettes to e-cigarettes. The strength of evidence and quality of the published studies overall is poor.

Our review has demonstrated that ENDS use is not causative for any harmful CVD outcomes and may be beneficial for hypertensive patients. Switching from cigarettes to e-cigarettes resulted in reduced exacerbations of COPD, with no evidence of increased risk of asthma, long-term respiratory harm or deterioration in lung function. Other health outcomes such as mental health, cancer and mortality have not been adequately studied to form a consensus. However, the findings of our review did not negate the consensus held by many that ENDS use is safer than the risks posed from smoking cigarettes.

Overall, our review found the research on ENDS use is not yet adequate to provide quantitative estimates about health risks. Consequently, the current body of evidence is inadequate for informing policy around tobacco harm reduction.

## Introduction

Smoking is the leading preventable cause of illness and premature death and one of the top causes of health inequalities, responsible for more than eight million deaths a year globally.^1^

The availability of tobacco harm reduction (THR) products has dramatically accelerated the reduction in smoking prevalence rate.^2^ Electronic nicotine delivery devices (ENDS), such as electronic cigarettes and vapes, are thought to be one of the most effective smoking cessation methods^3,4^ due to a combination of successful quit rates^5^ and their greater reach and accessibility compared with other smoking cessation methods.^2^

The prevalence of the use of ENDS is highest in the UK (6%) and the US (4-6%) compared with 1% the rest of Europe.^2^ The vast majority of regular ENDS users are previous or current smokers: in the UK over 99% of adult users and over 99.5% of adolescent users are former smokers;^2^ and in the US, 98.7% of adults aged 45 years or older and 60% of adults aged 18-24 years were former smokers.^6^

To date, there has been no clear consensus on the safety profile of ENDS and safety concerns have resulted in varying regulations and bans on their sale and use globally. The most widely used comparator for health risk is that of the Public Health England Report that estimated e-cigarettes to pose less than a 5% risk compared with conventional cigarettes.^7^ There have been no meta-analyses or systematic reviews to quantify the health risk posed by ENDS to date resulting in policy makers often using studies with flawed designs, or are animal, *in vitro* and *in silico* studies which may not translate to health outcomes in the real world.

The objective of this systematic review was to identify, narratively synthesize, assess the strength and quality of evidence and critically appraise studies that have reported health outcomes associated with use of ENDS.

## Methods

The aims of this study was to conduct a systematic review of the published literature on the health impact of ENDS products from 1^st^ January 2015 until 1^st^ February 2020. For the purpose of our study, ENDS included all electronic nicotine delivery devices but did not include heat-not-burn products. The study followed PRISMA guidelines for reporting of systematic reviews.^8^

### Search strategy and eligibility criteria

A literature search was conducted between 1^st^ October 2019 and 26^th^ February 2020 using the databases PubMed, Embase, Scopus and Google Scholar. Medical subject headings were used in the execution of PubMed searches. The search strategy encompassed two domains, including one for ENDS and related products, and one for health outcomes, including terms for cardiovascular disease (CVD), cancer, respiratory, mortality and ‘other’ health outcomes. Search results were restricted to English language reports, human studies and studies published since 2015, because most ENDS use has fallen within this period and because ENDS products available prior to 2015 have evolved considerably. The references of relevant reviews were manually searched for additional eligible citations. The detailed search strategy is provided in Appendix 1.

Search results were stored in Excel and de-duplicated before screening. The titles, abstracts and full texts of the search results were sequentially screened by two reviewers independently for inclusion using the eligibility criteria below, with disagreements resolved by blind review by a third reviewer.

Figure 1 shows the inclusion and exclusion criteria used.

**Figure 1.**
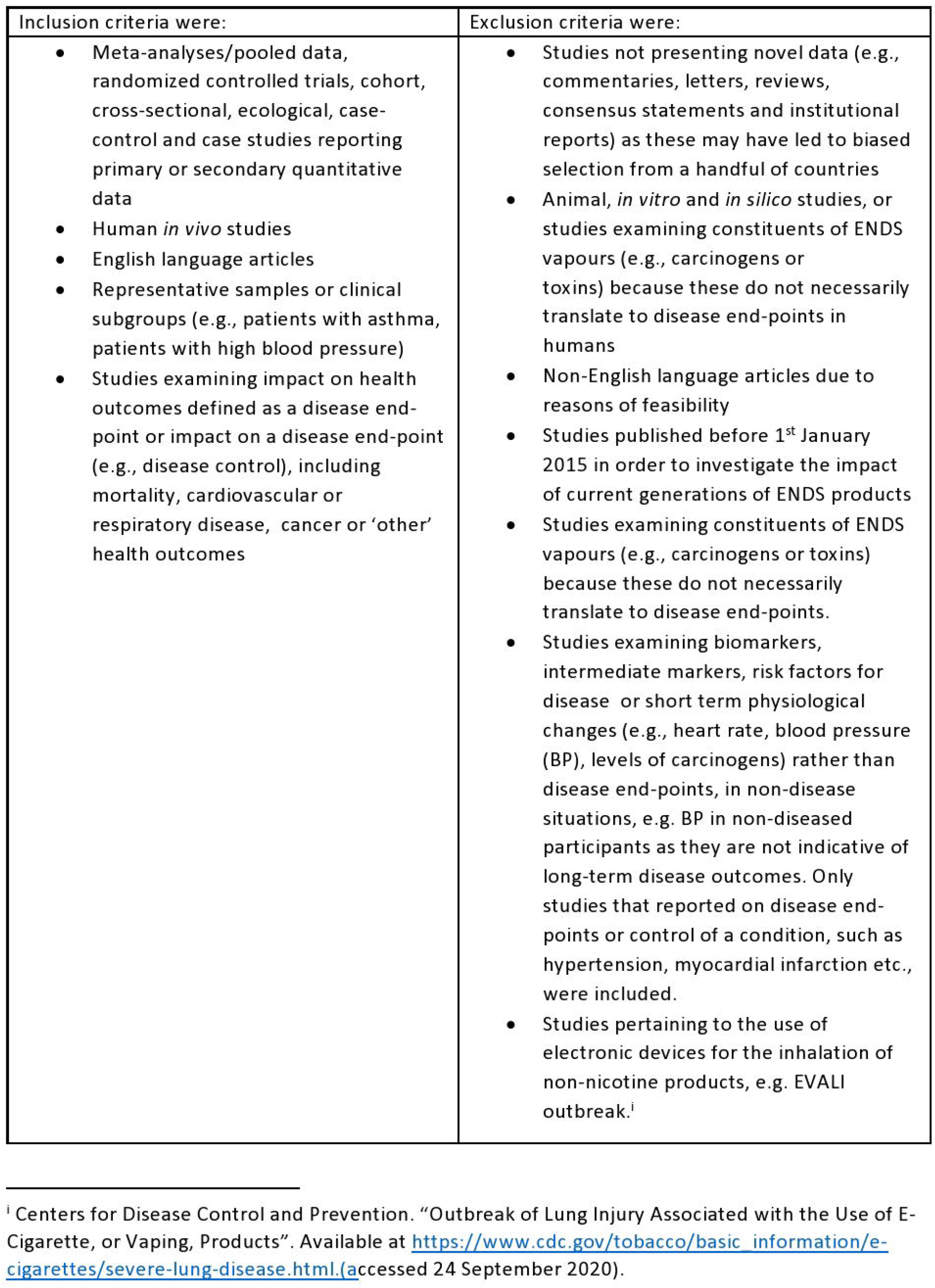
Inclusion and Exclusion Criteria.

Reasons for excluding studies were documented and are shown in figure 2.

**Figure 2.**
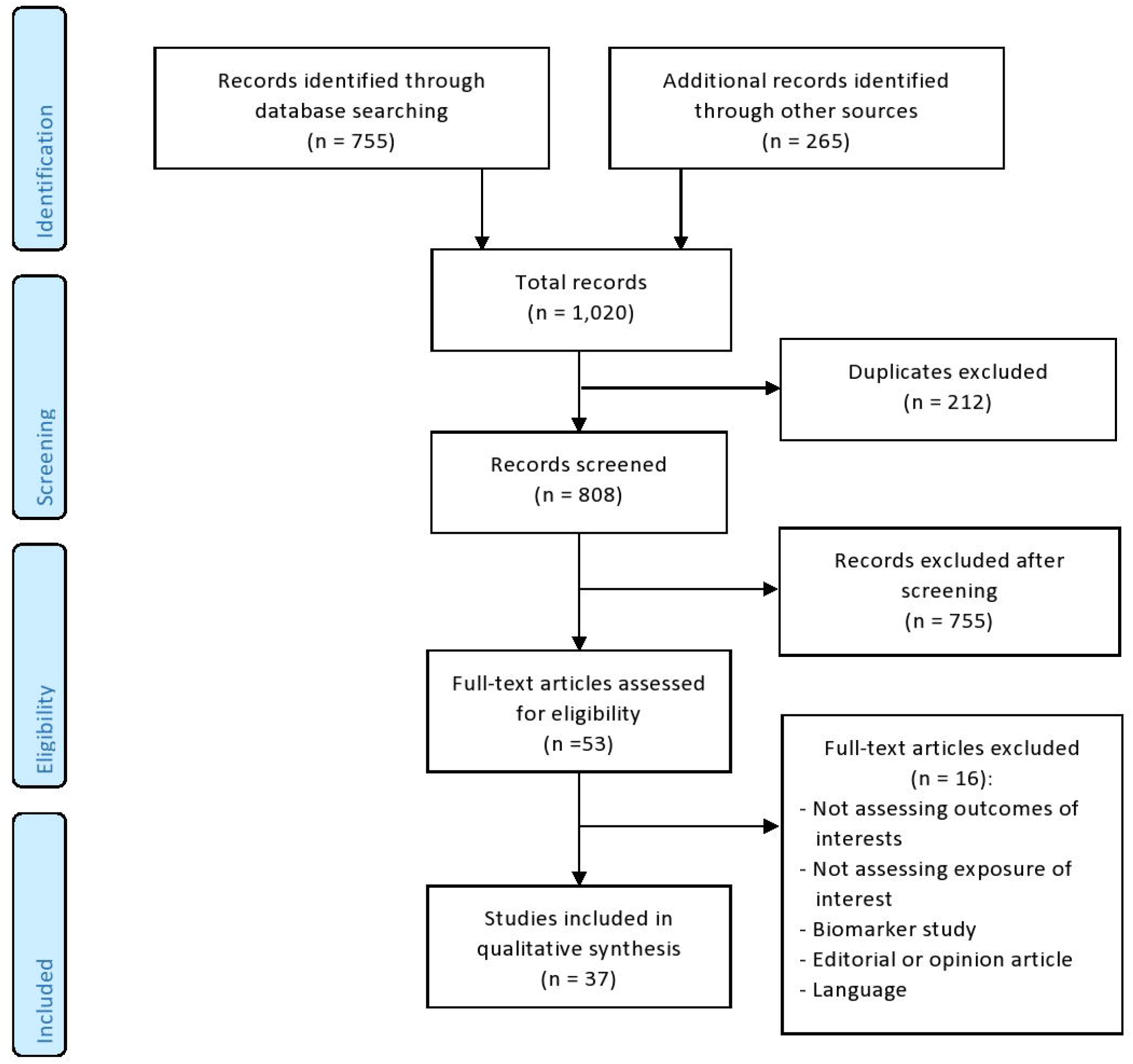
PRISMA flow chart of Included studies and Selection process.

### Data extraction and quality assessment

For all included studies, relevant data were extracted using a custom-designed table including author, year, country, aim, study design, sample size, participants, and relevant findings, including effect sizes and nature of impact on health outcomes. A category for the level of evidence was assigned using the Oxford Centre for Evidence Based Medicine framework.^9^ A similar approach was adopted to evaluate methodological quality, with each study assigned a quality rating of “good”, “fair” or “poor” utilizing the National Institutes for Health (NIH) Quality Assessment Tools.^10^ All data extraction and quality assessments were performed by two reviewers independently, with consensus reached with involvement of a third reviewer in cases of disagreement. Findings of all studies were independently read and re-read, coded, and organised into categories, which were then compared across studies to identify relationships and themes.

## Results

Thirty-seven studies were included in the review. Table 1 shows that the majority of studies were judged to be of low strength for levels of evidence, such as cross-sectional (24 studies, 65%), case-control (1 study, 2.7%) and case studies (6 studies, 16%). There were four (11%) cohort studies and one (2.7%) RCT. There was one (2.7%) MA/pooled study and no pooled results of randomized controlled trials (RCTs).

**Table 1.**
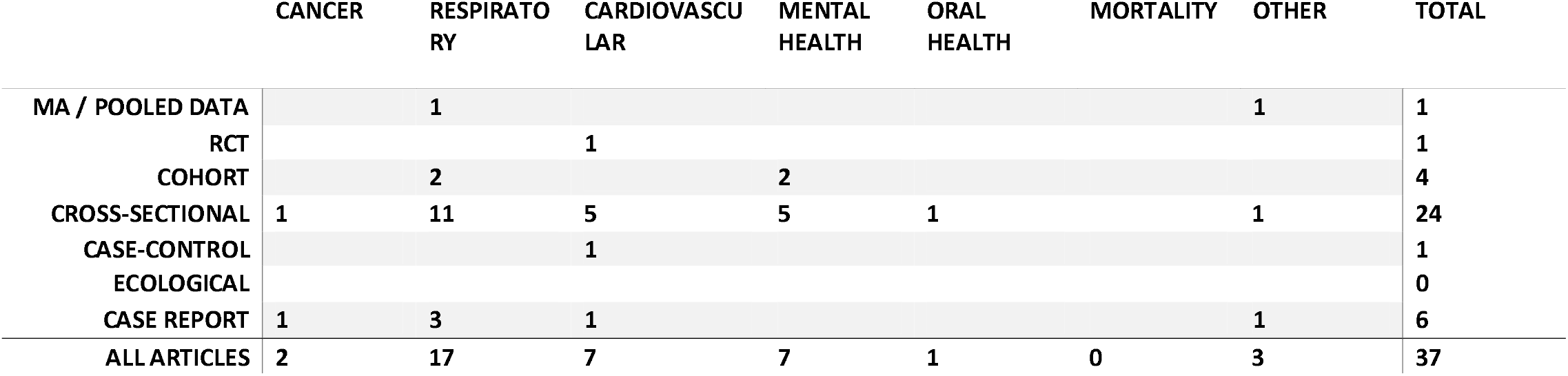
Number of Studies by Health Outcome and Study Design

Respiratory disorders made up 17 of the studies, followed by CVD (7), mental health (7) and one each on oral health, self-reported chronic health conditions, tonsillitis and nickel contact allergy.

Table 2 summarises that 27 (73%) of the studies examined and reported only on harms, eight (22%) on benefits, two (2%) reported on both harm and benefits. The one meta-analysis (MA)/pooled data study reported on harms, the one RCT was on benefits, and of studies investigating harms only, the majority (20, 74%) were cross-sectional, with 1 (4%) MA/pooled analysis, 1 (4%) cohort and five (19%) case-studies. Although few in number, studies investigating benefits tended to be of higher levels of evidence with one RCT, three cohort (including two studies examining both harms and benefits), four cross-sectional, one case-control and one case study.

**Table 2.** Study Design by Reporting of Harms and Benefits

|  | HARMS | BENEFITS | BOTH/NEUTRAL |
| --- | --- | --- | --- |
| META-ANALYSIS/POOLED DATA | 1 |  |  |
| RCT |  | 1 |  |
| COHORT | 1 | 1 | 2 |
| CROSS-SECTIONAL | 20 | 4 |  |
| ECOLOGICAL |  |  |  |
| CASE-CONTROL |  | 1 |  |
| CASE REPORT | 5 | 1 |  |
| ALL ARTICLES | 27 | 8 | 2 |

The studies reporting on benefits from ENDS included two studies on hypertension development and control, one on oral cancer development, two on COPD exacerbation, one on respiratory infections and one on depressive symptoms. A further two studies reported on both harms and benefits of ENDS, both on depressive symptoms.

Table 3 summarizes the quality ratings assigned to studies by health outcome. Raters one and two agreed on 32 out of 37 (94%) assessments of quality and level of evidence. “Poor” quality studies made up 20 (54%), “fair” made up nine (24%) and “good” made up eight (22%). Reasons for assigning poor quality ratings included insufficient follow-up for the outcome of interest to develop, inability to determine temporality and reverse causation, inadequate accounting for confounders and poor definitions of exposures and outcomes.

**Table 3.** Quality Ratings Assigned to studies by Health Outcome

| HEALTH OUTCOME | GOOD | FAIR | POOR |
| --- | --- | --- | --- |
| *ENDS* |  |  |  |
| MORTALITY | 0 | 0 | 0 |
| CVD | 2 (29%) | 2 (29%) | 3 (43%) |
| RESPIRATORY | 3 (18%) | 3 (18%) | 11 (65%) |
| CANCER | 0 (0%) | 2 (100%) | 0 (0%) |
| ORAL HEALTH | 0 (0%) | 0 (0%) | 1 (100%) |
| MENTAL HEALTH | 2 (29%) | 1 (14%) | 4 (57%) |
| OTHER | 1 (33%) | 1 (33%) | 1 (33%) |
| **TOTAL** | **8 (22%)** | **9 (24%)** | **20 (54%)** |

The characteristics of included studies, including study design, key outcomes, level of evidence and quality rating, are detailed in Table 4.

**Table 4.** Description of Studies, Level of Evidence and Quality for Health Outcomes from ENDS.

| **Cardiovascular** |  |  |  |  |  |  |  |
| --- | --- | --- | --- | --- | --- | --- | --- |
| Benefit | Farsalinos, et al. Effect of continuous smoking reduction and abstinence on blood pressure and heart rate in smokers switching to electronic cigarettes. | Italy | Prospective, double-blind, controlled, three-arm RCT on 145 hypertensive smokers switching to EC | Clinic measured SBP and HR | Reduced SBP (from 141 to 132 mmHg, $p<0.001$) in hypertensives switching to EC at 12 months; those continuing to smoke had no reduction. | 1B | Good |
| Harm | Alzahrani, et al. Association between electronic cigarette use and myocardial infarction. | United States | Cross-sectional survey on 69,04 subjects | Self-reported MI | Self-reported daily EC users more likely to report MI compared with never EC users (OR= 1.79; 95% CI: 1.20-2.66). Compared to never users of e-cigarettes and cigarettes, daily dual users of e-cigarettes and cigarettes were more likely to have an MI (OR=4.62) | 2C | Poor |
| Harm | Farsalinos, et al. Is e-cigarette use associated with coronary heart disease and myocardial infarction? | Italy | Cross-sectional survey in 2016 (n = 33,028) and 2017 (n = 26,742) | Self-reported MI and CHD | Self-reported daily EC use not associated with MI (OR= 1.35; 95% CI:0.80–2.27) compared with never EC use after accounting for dual use and former smoking; no association between EC use and CHD compared with never EC use (OR= 1.31; 95% CI:0.79-2.17) | 2C | Fair |
| Harm | Osei, et al. Association between e-cigarette use and cardiovascular disease among never and current combustible-cigarette smokers. | United States | Cross-sectional survey on 449,092 participants | CVD (defined as Self-reported CHD, MI, stroke) | Self-reported EC using never-smokers had no increased CVD (OR= 1.04; 95% CI: 0.63-1.72) or premature CVD (OR=1.01; 95% CI: 0.56-1.83) compared with never EC users; EC using former-smokers were more likely to report CVD (OR= 1.36; 95% CI= 1.18-1.56) and premature CVD (OR=1.45; 95% CI: 1.20-1.74) compared with never EC users. Dual smoking and EC use was associated with higher CVD (OR=1.36; 95% CI: 1.18-1.56). | 2C | Poor |
| Harm | Parekh, et al. Risk of Stroke With E-Cigarette and Combustible Cigarette Use in Young Adults. | United States | Cross-sectional survey on 161,529 participants aged 18–44 years | Self-reported stroke | Self-reported EC using never-smokers had no higher risk of stroke (OR=0.69, 95% CI: 0.34, 1.42) compared with nonsmokers; risk of stroke was lower for EC users compared with current exclusive smokers (OR=0.43, 95% CI: 0.20, 0.93). Current EC using former smokers had increased odds of stroke (OR=2.54; 95% CI: 1.16-5.56) compared with never -smokers. | 2C | Fair |
| Benefit | Polosa, et al. Blood pressure control in smokers with arterial hypertension who switched to electronic cigarettes. | Italy | Observational study of 89 hypertensive smokers who quit or reduced tobacco consumption by switching to EC. | Office SBP and DBP | A significant reduction in median SBP (from 140 to 130 mmHg; $p < 0.001$) and DBP (from 86-80 mmHg; $p = 0.006$) at 12-month follow-up in the exclusive EC group. No change in SBP or DBP seen in reduced cigarette consumption dual users at 12 months. | 3B | Good |
| **Respiratory** |  |  |  |  |  |  |  |
| Harm | Bowler, et al. Electronic cigarette use in US adults at risk for or with COPD: analysis from two observational cohorts. | United States | Pooled results from two cohort studies in 4,596 current or former smokers Aged 45–80 with, or at risk of, COPD | COPD respiratory symptoms or disease progression (GOLD criteria used to assess COPD spirometric severity) | Self-reported ever use of EC associated with 8% ($\pm 2\%$) increased prevalence of chronic bronchitis and (in 1 cohort) COPD exacerbations compared with never EC users ($p = 0.01$); after 5 years, no increase in progression of lung disease or decline in lung function (in one cohort). Adjusted for but not excluding current smokers. | 2A | Good |
| Benefit | Polosa, et al. Health effects in COPD smokers who switch | Italy | Prospective cohort study of 44 COPD smokers switching to | COPD exacerbations, post-bronchodilator lung function, CAT | Improvements in COPD exacerbation rates ($p=0.004$), CAT scores ($p=0.019$) and 6-minute walk distance ($p=0.001$) in EC users compared | 2B | Good |
|  | to electronic cigarettes: a retrospective-prospective 3-year follow-up. |  | EC use. | scores, 6-minute walking distance | with continued smokers after 36 months. |  |  |
| Harm | Lappas, et al. Short-term respiratory effects of e-cigarettes in healthy individuals and smokers with asthma. | Greece | Cohort study of 54 dual smokers (EC and smoking), 27 (50%) with mild asthma (MA), 27 (50%) no asthma, underwent a control session (no liquid, no resistor coil inside e-cigarette cartridge) and an experimental session of EC using standardized puffing settings. | Impulse oscillometry impedance (Z), resistance (R), reactance (X) and fractional exhaled nitric oxide (FeNO) were measured before and 0, 15 and 30 min after control and experimental sessions. | MA group exhibited higher baseline values and more prominent effect after EC use using standardized puffing sessions vs. healthy participants after EC use for respiratory system total impedance at 5 Hz (P = 0.022), respiratory system resistance at 5 Hz (P = 0.010) and respiratory system resistance at 10 Hz (P = 0.013). Fractional exhaled nitric oxide decreased significantly in both groups (P < 0.001) | 2B | Poor |
| Benefit | Polosa, et al. Evidence for harm reduction in COPD smokers who switch to electronic cigarettes. | Italy | Retrospective chart review with 12 and 24 month follow-up on 48 heavy smokers with COPD invited to switch to e-cigarettes | Verified COPD exacerbations in previous 12 months | Reduction in annual COPD exacerbations for heavy smokers with COPD switching to EC (mean 2.3 at baseline to 1.8; p=0.002) at 12 months and to 1.4 ;p< 0.001) at 24 months,: no change in COPD exacerbations for those not switching. | 2C | Good |
| Benefit | Miler, et al. Changes in the frequency of airway infections in smokers who switched to vaping: results of an online survey. | Germany | Cross-sectional survey of 914 smokers who switched to vaping for at least two months. | Self-reported respiratory infections (e.g., common cold) | Among those who switched to EC, 66% (95% CI=62.9-69.0) reported improvement in respiratory infections, 29% reported no change, 5% reported worsening. | 2C | Poor |
| Harm | Bhatta, et al. Association of e-cigarette use with respiratory disease among adults: a longitudinal analysis. | United States | Cross-sectional survey of 705,159 participants | Self-reported chronic bronchitis, emphysema, COPD | Among never smokers, current self-reported EC use associated with chronic bronchitis, emphysema and COPD compared with never EC users (OR=1.75, 95% CI: 1.25-2.45); daily EC use had higher odds (OR=2.64, 95% CI:1.43, 4.89). | 2C | Poor |
| Harm | Cho, et al. Association between electronic cigarette use and asthma among high school students in South Korea.<br>**There are no sources in the current document.** | South Korea | Cross-sectional survey of 35,904 high school students | Self-reported asthma diagnosis | Among self-reported never smokers, current EC use associated with asthma (OR=2.74; 95% CI: 1.30–5.78) compared with never EC users. | 2C | Poor |
| Harm | Choi, K., & Bernat, D. (2016). E-cigarette use among Florida youth with and without asthma. | United States | Cross-sectional survey of 36,085 high school students | Self-reported asthma and asthma attack | Among those with asthma, self-reported past 30-day EC use (any quantity) associated with asthma attacks (OR=1.78, 95% CI: 1.20-2.64) in the past 12 months compared with non EC users in past 30 days (adjusted for days smoked cigarettes in the past 30 days but smokers not excluded) | 2C | Poor |
| Harm | Kim, et al. Active, passive, and electronic cigarette smoking is associated with asthma in adolescents. | South Korea | Cross-sectional survey of 216,056 adolescents aged 12-18 years | Self-reported asthma | Self-reported EC use group associated with higher prevalence of asthma (OR=1.13; 95%CI: 1.01–1.26) compared with never EC users (adjusting for active, passive cigarette use); greater use of e-cigarettes associated with asthma, 1-5 days/month (OR=1.39; 95% CI: 1.19-1.61), 6-19 days/month (OR=1.31; 95% CI: 1.08-1.61) and >20 days/month (OR=1.58; 95% CI: 1.40-1.78) compared with never EC use. | 2C | Poor |
| Harm | Osei, et al. Association Between E-Cigarette Use and Chronic Obstructive Pulmonary Disease by Smoking Status: Behavioral Risk Factor Surveillance System 2016 and 2017. | United States | Cross-sectional survey on 5,454 participants | Self-reported COPD diagnosis | Self-reported non-current smokers using EC associated with a COPD diagnosis (OR=2.94, 95% CI: 1.73–4.99) compared with non-EC use. Compared with never smokers who never used EC, dual users (smoking and EC) had the highest odds of COPD (OR=6.89, 95% CI=6.29, 7.55). Former smoking was not excluded or accounted for. | 2C | Poor |
| Harm | Perez, et al. Adult e-cigarettes use associated with a self-reported diagnosis of COPD. | United States | Cross-sectional survey of 32,320 adults and adolescents aged 12–17 years | Self-reported COPD diagnosis | Self-reported EC users had greater odds of COPD than non- EC users (OR=1.43, 95% CI: 1.12–1.85) in adults and children combined. | 2C | Poor |
| Harm | Schweitzer, et al. E-cigarette use and asthma in a multiethnic sample of adolescents. | United States | Cross-sectional survey of 6,082 adolescents | Self-reported asthma diagnosis | Current self-reported EC use associated with asthma (OR=1.48; 95% CI: 1.24–1.78) and with previous asthma (OR=1.20; 95% CI: 1.00–1.44) compared with never EC use, (controlling for but not excluding current cigarette smoking, or former smoking). | 2C | Poor |
| Harm | Wills et al. E-cigarette use and respiratory disorder in an adult sample. | United States | Cross-sectional random-dial telephone survey on 8,087 adults | Self-reported asthma or COPD diagnosis | Self-reported ever EC use associated with asthma in current non-smokers (OR = 1.33, 95% CI: 1.00–1.77, p<.05) but not in smokers (OR=0.92, 95% CI: 0.73–1.15, EC use was not associated with COPD in current non-smokers (OR = 2.98, 95% CI: 1.51–5.88, p < .01) or in current smokers (OR = 1.29, 95%CI 0.94 –1.77). There was no significant difference in risk of asthma among dual users compared with sole EC users (OR=1.00; 95%CI: 0.73-1.35) or smokers (OR=0.99; 95%CI: 0.80-1.22). There was increased risk of COPD in smokers (OR=2.98; 95%CI: 2.34-3.78), EC users (OR=2.58; 95%CI: 1.36-4.89) and dual users (OR=3.92; 95%CI: 2.82-5.44) compared with never smokers who never used EC. Ever EC use included any quantity ever used. | 2C | Fair |
| Harm | Xie, et al. Use of Electronic Cigarettes and Self-Reported Chronic Obstructive Pulmonary Disease Diagnosis in Adults. | United States | Cross-sectional survey of 887,182 participants | Self-reported COPD diagnosis | Self-reported current vapers who never smoked more likely to self-report COPD (OR=1.47; 95% CI: 1.01, 2.12) compared with never smokers (smoked less than 100 cigarettes, not currently vaping). | 2C | Fair |
| Harm | Sommerfeld et al. Hypersensitivity pneumonitis and acute respiratory distress syndrome from e-cigarette use. | United States | Case study, 18 year old woman with dyspnea, cough, and pleuritic chest pain after e-cigarette use | Hypersensitivity Pneumonitis and acute respiratory distress syndrome | Case study of single EC user developing sensitivity pneumonitis. Did not report on comorbidities or smoking | 4 | Fair |
| Harm | Khan, et al. Organizing pneumonia related to electronic cigarette use: a case report and review of literature. | United States | Case study, 40-year-old female patient | Organising pneumonia | Single case study of organizing pneumonia, exclusion of other drug use and comorbidities not mentioned. | 4 | Poor |
| Harm | Carter, et al. Life-threatening vesicular bronchial injury requiring veno-venous extracorporeal membrane oxygenation rescue in an electronic nicotine delivery system user. | United States | Case study, 35-year-old female presented to emergency department with chest pain and dyspnea | Vesicular Bronchial Injury | Case study showed vesicular bronchial injury in an EC user. Patient had CVD and other comorbidities and was a former smoker. | 4 | Poor |
| **Cancer** |  |  |  |  |  |  |  |
| Benefit | Franco, et al. Electronic cigarette: role in the primary prevention of oral cavity cancer. | Italy | Cross-sectional survey on 65 previous smokers (from outpatient center), e-cigarette smokers (from monthly prevention campaigns), and nonsmokers (from university medical and paramedical staff) | Total number of oral mucosa pre-cancerous (micronucleated) cells from cytologic examination | Self-reported EC users had lower micronuclei compared with smokers based on micronucleated cells/1000 cells (P = 0.001) and micronuclei/1000 cells (P = 0.004) | 2C | Fair |
| Harm | Nguyen, et al. Oral carcinoma associated with chronic use of electronic cigarettes. | United States, Vietnam | Case study of 2 subjects | Oral carcinoma | Two cases of oral carcinoma associated with 13-year use of EC. Description of other risks not detailed eg smoking. | 4 | Fair |
| **Oral Health** |  |  |  |  |  |  |  |
| Harm | Akinkugbe, et al. Cigarettes, E-cigarettes, and Adolescents' Oral Health: Findings from the Population Assessment of Tobacco and Health (PATH) Study. | United States | Cross-sectional study on 13,650 adolescents aged 12 to 17 years | Dental problems (cavities, gum disease or dental stains) | No relationship between self-reported EC use and self-reported dental problems, including among current eEC users (OR=1.11; 95% CI: 0.79-1.55) or ever users (OR=1.12 95% CI: 0.90-1.38) compared with never cigarette or EC users. | 2C | Poor |
| **Mental Health** |  |  |  |  |  |  |  |
| Harms and Benefits | Bandiera, et al. Depressive symptoms predict current e-cigarette use among college students in Texas. | United States | Cohort study of 5,445 college students (18–29 year olds) with 6-month and 1-year follow-ups | Self-reported depressive symptoms | Correlation between depressive symptoms and self-reported EC use was significant at baseline ($\beta = .05$; $p < .01$), however, EC use did not predict higher depressive symptoms at 6-months or 1-year follow-up. | 2B | Good |
| Neutral | Lechner, et al. Bi-directional associations of electronic and combustible cigarette use onset patterns with depressive symptoms in adolescents. | United States | Cohort study of 347 adolescents assessed at baseline, 6- and 12-month follow-up | Self-reported depressive symptoms | Self-reported EC use over previous 12-months associated with greater rate of increase in depressive symptoms over time ($b = 1.272$, $SE = 0.513$, $p = 0.01$) compared with never EC use. Higher frequency of EC use was associated with higher depressive symptoms at 12 months among sustained users ($B = 1.611$, $p = 0.04$). | 2B | Good |
| Benefit | Dahal, et al. Smoking Cessation and Improvement in Mental Health Outcomes: Do People Who Quit Smoking by Switching to Electronic Cigarettes Experience Improvement in Mental Health? | Canada | Cross-sectional survey on 52,956 participants | Self-reported depressive symptoms | Self-reported EC use (any quantity) who were never smokers had higher depressive symptoms ($\geq 10$ on CES-D 10) compared with never EC users ($OR=2.46$; 95%CI: 1.82-3.33). Former smokers who used ECs had higher depressive symptoms compared with never smokers ($OR=4.19$; 95%CI: 2.47-7.11). Former smokers who did not use EC had elevated risk of depressive symptoms as well ($OR=1.41$ (95% CI: 1.19 to 1.68) compared to never smokers. EC use included any quantity including experimental use. | 2C | Poor |
| Harm | Chadi, et al. Depressive Symptoms and Suicidality in Adolescents Using e-Cigarettes and Marijuana: A | United States | Cross-sectional survey of 26,821 high school students | Self-reported depressive symptoms and suicidal ideation | Self-reported EC use associated with higher odds of suicidal ideation in past 12 months ($OR=1.23$; 95%CI: 1.03-1.47) and depressive symptoms ($OR=1.37$; 95%CI: 1.19-1.57) compared with never EC users, adjusted for current smoking (but former and current | 2C | Poor |
|  | Secondary Data Analysis From the Youth Risk Behavior Survey. |  |  |  | smokers were not excluded). No use of validated scores to obtain outcomes. |  |  |
| Harm | Grant, et al. E-cigarette use (vaping) is associated with illicit drug use, mental health problems, and impulsivity in university students. | United States | Cross-sectional survey of 3,572 college and graduate school students | Self-reported mental health issues on PHQ9 scale, self-reported diagnosis of ADHD (Y/N), PTSD (PC-PTSD score), gambling disorder (Y/N), anxiety (GAD-7 score), trait impulsivity plus compulsivity, academic impairments | Self-reported EC use associated with mental health issues, including PHQ-9 score $\geq 10$ (Cramer's $V=.044$; $p=0.052$), ADHD (Cramer's $V=.073$; $p<.001$), PTSD (PC-PTSD score $\geq 3$; Cramer's $V=.064$; $p<.002$), gambling disorder (Cramer's $V=.081$, $p<.001$) and anxiety (GAD-7 $> 10$; Cramer's $V=.066$; $p<.001$). They were also more likely to report low self-esteem (Cramer's $V=0.63$; $p=.002$), and endorse traits of impulsivity (attentional: cohen's $d=.421$; $p<.001$), but not compulsivity (cohen's $d=0.532$; $p=.043$). Did not control for cigarette use. Participation rate of 38% so sample bias possible. No definition of EC use provided. | 2C | Fair |
| Harm | King, et al. Tobacco product use and mental health status among young adults. | United States | Cross-sectional survey of 2,370 college students | Self-reported depression (higher score, greater depression), stress (higher score, greater perceived stress), mental health diagnosis | Self-reported EC use associated with higher depression score (OR=1.04; 95% CI: 1.01-1.08) compared with never EC use, controlling for 30-day cigarette use. EC use was associated with higher stress score (OR=1.03 95% CI: 1.00-1.05) compared with never EC use, controlling for 30-day cigarette use. Dual use but not former smoking was accounted for. | 2C | Fair |
| Harm | Pham, et al. Electronic cigarette use and mental health: A Canadian population-based study. |  | Cross-sectional survey of 53,050 participants | Self-reported depressive symptoms, mood and anxiety, mental health, suicidal thoughts, binge drinking | Among female non-smokers, self-reported EC users had increased mood disorders (OR=1.9; 95% CI: 1.2–3.0) and anxiety disorders (OR=1.9; 95% CI: 1.1-3.2) compared with non- EC users. Female current EC use was associated with mood (OR=1.9 (95%CI: 1.4–2.6) and anxiety (OR=2.6 (95% CI: 1.9–3.6)) disorders compared | 2C | Poor |
|  |  | Canada |  |  | with non EC use. Among male non-smokers, self-reported EC users had increased mood disorders (OR=1.6; 95% CI: 1.0-2.7) compared with non-EC users. Among male smokers, EC use was not associated with mood disorders (OR=1.4 (95%CI: 0.9–2.3). EC use was defined as any quantity within the last 3 months, including experimental use. |  |  |
| **Other** |  |  |  |  |  |  |  |
| Harm | Lanza, et al. Obesity and cigarette smoking: Extending the link to e-cigarette/vaping use. | United States | Cross-sectional survey (convenience sample) of 452 participants | Self-reported BMI | Obese (BMI $\geq 25\text{Kg/m}^2$) participants had higher likelihood of belonging to self-reported Cigarette/EC / Tobacco class compared with the High Substance Use ($\beta = 1.48$, OR = 4.40, $p < .05$) and Risky Alcohol Use ($\beta = 1.94$, OR = 6.97, $p < .05$) classes; higher likelihood of being classified into the cigarette/ electronic tobacco class compared to the low substance use class not significant.<br>No detail of definitions for EC use. | 2C | Poor |
| Benefit | Miler, et al. Resolution of recurrent tonsillitis in a non-smoker who became a vaper. A case study and new hypothesis. | United Kingdom | Case study of a never-smoker who vapes, with a history of recurrent, chronic tonsillitis | Exacerbations of tonsillitis | After 8 months of vaping, the patient reported absence of exacerbations of tonsillitis, and marked improvement in Tonsillitis. The study did not mention any other comorbidities or exhaustively account for all confounders | 4 | Fair |
| Harm | Maridet, et al. The electronic cigarette: the new source of nickel contact allergy of the 21st century? | France | Case study on a 52-year old woman | Clinically-determined erythematous, scaly dermatitis | The patient was diagnosed with nickel contact dermatitis associated with the use of an electronic cigarette. The articles also discussed the literature on nickel content in different brands of ECs. | 4 | Good |

### Overall results for health outcomes by category

#### CVD Outcomes

Two studies, an RCT and observational study both rated as being of “high” quality, reported on improvements to the control of BP in hypertensive patients, finding reductions in systolic blood pressure (SBP) by 9-10mmHg, and diastolic BP (DBP) by 6mmHg.^11,12^

Three studies reported on the association between CVD (including acute myocardial infarction [MI]) and the use of ENDS. Two large cross-sectional surveys on approximately 0.5 million^13^ and 60,000^14^ subjects found that users of ENDS had no increase in MI, coronary heart disease (CHD), premature CVD or CVD compared with never smokers. However, former smokers who used ENDS did have more CVD (OR 1.4) and premature CVD (OR 1.5) than never smokers in one of the studies.^13^ Dual users experienced higher CVD (OR 1.36) compared with those who were current smokers not using ENDS.13 A further study that did not account for former smokers or dual users, or for temporality and reverse causation, found users of ENDS to have increased risk of MI (OR 1.8).^15^

A large cross-sectional study investigating stroke found no excess risk in users of ENDS in never smokers.^16^ The use of ENDS in ex-smokers was associated with a higher risk of stroke (OR 2.5) compared with never smokers.^16^

#### Respiratory Outcomes

The majority of respiratory outcomes were on the development or exacerbation of chronic obstructive pulmonary disease (COPD) in adults or asthma in adolescents. A few further studies reported on other conditions such as rates of respiratory infection.

The most rigorous studies reporting on COPD were a pooled study of two cohorts,^17^ and an interventional study over 12 months,^11^ with further follow up over three years.^18^ Those studies found that COPD exacerbations reduced in frequency in heavy smokers switching to e-cigarettes from 2.3 to 1.4 annually,^18^ and improvements in verified COPD Assessment Test (CAT) score, walking distance and continued reductions in COPD exacerbations after three years.^18^ A study pooling findings from two cohort studies,^17^ without excluding current smokers, reported e-cigarette users to have 8% higher prevalence of chronic bronchitis and COPD exacerbations in one of the included cohort studies. After five years of follow-up, no increased progression of lung disease or decline in lung function was seen in e-cigarette users.^17^ Current and former smoking was adjusted for but not excluded.

Five cross-sectional studies^13,19,20,21,22^ investigated the association between e-cigarette use and COPD. In one of these studies,^20^ 85% of the sample did not fall into the age-risk category (age over 55 years) for COPD.^23^ One of the cross-sectional studies on a sample of almost 900,000 never-smokers showed an association (OR 1.5) between e-cigarette use and self-reported COPD compared with non-e-cigarette use.^22^ Another study that segmented never and current smokers only found an association between e-cigarettes and COPD in smokers (OR 1.3) and not in never-smokers (OR 0.9).^21^

Six studies investigated the development or control of asthma.^21,24,25,26,27,28^ An experimental study showed that following e-cigarette use, respiratory system resistance and impedance were impacted up to 30 minutes post, but fractional exhaled nitric oxide did not differ between asthmatics and non-asthmatics.^28^ Five of the six studies were cross-sectional in design and several relied on children and adolescents self-reporting on e-cigarette use and a diagnosis of asthma in schools and other educational facilities. The definitions of e-cigarette users included experimental and one-time use of e-cigarettes in some studies.^24,25,26,28^ One of the cross-sectional studies^21^ reported separately for never smokers and smokers, and found e-cigarette use to be associated with a higher rate of asthma in smokers (OR 1.3) but not in non-smokers (OR 0.9). Another large cross-sectional study^24^ reported an association of e-cigarette use in never smoking adolescents with a self-reported diagnosis of asthma (OR 2.7). The remaining studies reported associations between e-cigarette use and asthma, with OR’s ranging between 1.1 and 1.8.^21,26,28^

In a cross-sectional study on 914 smokers who switched to e-cigarettes, 66% reported reductions in the frequency of respiratory infections and 6% reported worsening.^29^ Single case studies reported on acute hypoxaemic respiratory failure and organizing pneumonia; organizing pneumonia; sensitivity pneumonitis and vesicular bronchial injury, but none specifically excluded other causes such as dual use, former smoking, other drug use or comorbidities.

#### Cancer

A small cross-sectional study demonstrated lower numbers of oral cancerous cells (50%) and cellular changes (33%) in e-cigarette users who were never smokers compared with smokers (p=0.001).^30^ The other was a case study of two individuals who developed oral cancer after 13 years of e-cigarette use. Other risk factors such as smoking were not mentioned as having been excluded.

#### Mental Health

Seven studies reported on the association between ENDS use and mental health disease. Of two cohort studies,^31,32^ the first found that those with depressive symptoms were more likely to take up e-cigarette use at six months (beta coefficients 0.06 & 0.08), but at 12 month follow-up, they did not have more depressive symptoms than non e-cigarette users.^31^ Another cohort study^32^ found a greater increase in depressive symptoms in e-cigarette users after 12 months (beta=1.27, p0.01) compared with non e-cigarette users, with a positive dose-response effect.

Four cross-sectional studies^33,34,35,36^ reported a positive association between e-cigarette use and self-reported depressive symptoms with wide-ranging ORs from 1.03 to 4.2.

Another cross-sectional study^50^ found an association between e-cigarette use and attention-deficit hyperactivity disorder (ADHD; V=.073; p<.001), post-traumatic stress disorder (PTSD) (V=.064; p=<002), gambling disorder (V=.081, p<.001), anxiety (V=.066; p<.001), low self-esteem (V=0.63; p=.002) and impulsivity traits (cohen’s d=.421; p<.001). The study did not control for cigarette use, had a participation rate of only 38% with potential sample bias and did not state a definition for e-cigarette use, in addition to not accounting for reverse causation.

#### Oral Health

One study reported on dental health outcomes;^37^ a cross-sectional study that reported no association with self-reported dental health issues in e-cigarette users compared with never smokers.

#### Other health outcomes

One cross-sectional study reported an association between e-cigarette use and obesity (OR 4.4, p < .05) and alcohol abuse (OR 7.0, p < .05).^38^ There were two single case studies of e-cigarette use being linked to improvement of recurrent tonsillitis^29^ and occurrence of nickel contact allergy.^39^

#### Mortality

No studies were found that investigated mortality related to the use of ENDS.

## Discussion

In order to determine the net health impact of ENDs, the benefits from quitting smoking must be weighed against any harms (or benefits) from the use of ENDS. The wider impacts from the use of ENDS on society, such as new uptake in never smokers and nicotine addiction, must also be factored in, which are outside of the scope of this review.

This is the first article to systematically review the health outcomes from ENDS. Over the five-year period, 37 studies were identified. We found that studies tended to focus on the negative health impacts from ENDS, with the benefits of switching from cigarettes to ENDS being an uncommon outcome measure. Evidence of significant harms to health outcomes from ENDS was lacking from our review, with the majority of study designs being unable to rigorously establish causation. In the handful of studies that were of adequately rigorous design, no causation has been established between the use of ENDS and negative health outcomes. There is some evidence of positive health outcomes in those switching from cigarettes to e-cigarettes but further studies would be required to replicate the findings.

### Levels of evidence, quality and study design

The sample size of studies was not a common study flaw found in this review. Due to the nature of the research question on self-reported smoking and use of ENDS, numerous studies used pre-existing survey data with 887,182 participants in the largest study. However, the large size of studies did not reflect their quality which was often poor.

There were no studies rated above 2a for level of evidence, i.e. there were no MAs or pooled study designs on RCTs. Experimental rather than observational designs are desirable so that confounders and biases can be adequately accounted for but the vast majority of studies (97%) in our review were observational. There was only one interventional study investigating benefits from switching from smoking to e-cigarettes. The low number of RCTs in this review reflects the difficulty of conducting interventional THR studies in real world settings. There were few cohort studies, with only one study on harms, one on benefits and two exploring both harms and benefits.

Cross sectional studies were predominant (41%), without accounting for temporality and reverse causation, which is particularly relevant here as the majority of ENDS users are current or former smokers.^1,5,40^ Furthermore, those with smoking-related medical conditions such as asthma, COPD and CVD are more likely to switch to ENDS in order to quit smoking.^41^ Without accounting for temporality of the exposure and outcome, as well as former smoking status, many study findings are inadequate for causal inferences.

We considered a MA of studies included in our review to be inappropriate, partly due to the common methodological flaws highlighted above and the vast heterogeneity between studies, for example in the definitions used for the exposure variable of ENDS use, and with regards to accounting for dual use, former use, duration and quantity of use. Our review also found re-use of the same surveys and databases for several separate studies.^13,42,43^

Included studies were predominantly rated as being of poor quality. Studies that examined benefits to health outcomes had a relatively higher number of fair or good quality studies (6, 75%) compared with those on harms alone (9, 33%).

### Definition of Exposure

The definitions used by studies for smoking and use of ENDS varied tremendously. The vast majority of studies included here relied on self-reported data for smoking and use of ENDS, which is known to underestimate the true prevalence of smoking.^44^ Particularly problematic were several studies that asked children and adolescents in educational settings to self-report their use of cigarettes and ENDS which are usually prohibited. Furthermore, these studies used self-reported information from adolescents on health outcomes such as asthma and depression which may also be unreliable.^45^

Health outcomes from smoking cigarettes are dose-dependent.^46^ Similarly, it is likely that health outcomes for ENDS are also dose-dependent, yet the majority of studies failed to quantify this in their definitions for exposure. Studies with poor definitions of smoking failed to account for quantity, duration since quitting and duration of ENDS use, dual and former use of cigarettes and ENDS. ^12^,13,^16,17,33,36^ Studies using data from the Population Assessment of Tobacco Health (PATH) and Behavioral Risk Factor Surveillance System datasets13^,16,20,22,47,48^ and others,^34,31^ respondents who ever used a cigarette, other tobacco product or ENDS, even once or twice, were regarded as former or current users. Those having ever experimented with cigarettes or e-cigarettes were therefore regarded in the same category as heavy smokers or daily e-cigarette users.

Standard definitions exist for smoking, such as smoking 100 or more cigarettes, smoking at least one cigarette daily for 12 months or cigarette pack years (CPY) which accounts for both quantity and duration of smoking, both of which impact health outcomes.^49^ Similar approaches should be used to quantify use of ENDS. Some good definitions of smoking and ENDS use were seen that accounted for quantity, duration, dual and former use.^11,32,22^

### Definitions of Outcomes

Both exposures and outcomes were self-reported in the majority of studies, and only 14 (38%) of studies utilized verified health outcomes data. Self-reporting of outcomes is known to be unreliable and prone to bias in some situations. Particularly problematic in this review were several studies that asked children and adolescents in educational settings to self-report on asthma and depressive symptoms both of which could have led to subjective and inaccurate responses.^24,25,26,28,50,51^

### Accounting for Smoking Status

One of the major design flaws of the included studies was a failure to account for current, former and dual use of cigarettes^52,53,54^ and in doing so, ignoring the known evidence that the majority of ENDS users do so for the purpose of quitting or cutting down on cigarette smoking.^52,53,54^ Several studies compared health risks for ENDS users with those of never smokers without accounting for former smoking in the ENDS users, such that they were making a comparison of predominantly ex-smoking ENDS users with never smokers, rather than exclusively reporting the impact of ENDS use. A more meaningful comparisons in this regard would be between exclusive ENDS users who were never smokers against non-ENDS users who were never smokers. In order to quantify the benefit from switching, former smokers who now exclusively used ENDS should be compared with current smokers, accounting also for the duration of switching, duration of smoking and the quantity of cigarettes smoked. These aspects of study design are required to look at causation rather than just correlation and to account for biases that were almost always overlooked in the studies included in the review.

Cigarette smokers often transition to ENDS rather than switching immediately, with 70% of e-cigarette users reporting dual use.^55^ This review found that studies do not routinely account for dual use when investigating risk from ENDS with the consequence of attributing health outcomes to the use of ENDS when they may instead result from smoking cigarettes.

### Temporality and Reverse Causation

A crucial and common methodological flaw was the failure to account for temporality and reverse causation as was seen in the 41% of studies included in our review that were cross-sectional in design, in view of the fact that the majority of adult and adolescent users are previous or current smokers.^2,6^ Furthermore, some of the health outcomes such as COPD and CVD can take up to decades to develop. Cross-sectional studies in current or former smokers cannot be used to establish temporal precedence as was reported in several studies, one of which has since been retracted.^42^ Studies reporting on mental health in particular failed to account for reverse causation.

### Publication Bias

The ratio of studies on harm versus benefits was high with only eight studies out of 37 investigating potential health benefits from use of ENDS compared with cigarettes, and two investigating both harms and benefits. It is possible that there has been a tendency to more frequently report on harmful health outcomes rather than neutral or beneficial one. Indeed, we observed that some studies where the primary research question was to investigate harmful impact on health outcomes did not explicitly report findings that were either neutral or beneficial to health outcomes in the abstract and text of the article, focusing instead on the findings related to harms to health outcomes.^22^ Future studies will need to prioritise an exploration of both potential harms and benefits.

### Health Outcomes

The majority of health outcomes fell under the categories of respiratory (46%), CVD (22%), cancer (5%), oral health (3%) and mental health (19%). Other health outcomes included nickel contact dermatitis (one study), obesity (one study) and tonsillitis (one study).

### Mortality

In the US, the CDC report that overall mortality among both male and female smokers is three times higher than that among similar people who never smoked.^56,57^ The major causes of excess mortality among smokers include cancer, respiratory disease and CVD,^58,59,60^ and some types of smokeless tobacco are known to cause cancer and related mortality.^56^ Furthermore, quitting smoking before the age of 40 has been shown to reduce the risk of dying from smoking-related disease by about 90%.^2,58^

It is surprising, therefore, that this is not reflected in the focus of research on harms from ENDS, with no studies identified in the last five years looking at the association between ENDS and mortality. Whilst this may be partly due to the relatively recent availability of ENDS, it would be feasible to study mortality as an outcome in studies of high-risk groups such as CVD patients.

### Cardiovascular Disease

There is an extensive body of evidence showing that smoking tobacco is causally related to almost all major forms of CVD,^61^ including accelerated atherosclerosis and an increased risk of acute MI, stroke, peripheral arterial disease (PAD), aortic aneurysm, sudden death and many risk factors for CHD. Among adults 55–74 years of age, an estimated two-thirds of CHD deaths are attributable to smoking^62^ and the benefits of quitting smoking on reduced risk for CHD and CVD mortality have been well documented.^63,64,65,66,67^

We had expected to see more robust studies on the impact of switching from cigarette smoking to ENDS. The recent availability of ENDS may be partly responsible although other diseases such as COPD have been reported within the same timelines.

Our review has found that use of ENDS products has not been shown to be causative for any CVD outcomes and has been shown to be beneficial for patients with hypertension. Further interrogation using longitudinal study design and longer follow-up are needed to definitively confirm the lack of harm.

### Respiratory Disease

The US Surgeon General’s report in 1964 reported smoking as the most important cause of COPD^68^ with a relative risk of dying of approximately 26 for men and 22 for women.^62^ Quitting of smoking early on in the COPD disease timeline is associated with reductions in morbidity and mortality.^69^ Smoking has also been shown to increase the risk of developing asthma in adolescents, and of triggering asthma attacks and worsening outcomes of attacks.^57,70^ Other lung disorders that are causally linked with smoking include tuberculosis (TB) and idiopathic pulmonary fibrosis.^57^

Cross sectional designs are particularly problematic as the development of COPD usually takes several decades.^71^ Also, patients developing COPD would be advised to reduce or quit smoking by their physician and may be more likely to take up quit aids such as e-cigarettes.

Although the results in our review are mixed, the only studies judged to be of rigorous design (i.e. accounting for temporality, and former and current smoking) suggest that switching from cigarettes to e-cigarettes results in a reduction in exacerbations of COPD, with no evidence of long-term deterioration in lung function. The best evidence found no increased risk of asthma in ENDS users who were never smokers. There is a suggestion of short-term respiratory function changes in asthmatics using ENDS, but we do not know if these would translate to long-term impact.

### Cancer

The link between cancer and cigarette smoking has been extensively studied since the landmark study to report unequivocally that smoking impacts on rates of lung cancer in doctors who smoked.^72^ Other smoking-related cancers are of the mouth, throat, nose, sinuses, oesophagus, bladder, kidney, ureter, pancreas, stomach, liver, cervix and ovary, bowel and acute myeloid leukaemia.^73^

Only two studies on cancer were identified in this review. The association of e-cigarettes in the causation of cancer has not been explored in clinical studies to any extent, which may in part be due to the lack of a plausible biological pathway.

### Oral Health

Oral cancer is the eleventh most common cancer worldwide.^74^ The vast majority of studies reporting on oral health were on periodontal issues rather than health outcomes hence were not included. Oral health issues for ENDS has not been adequately studied to form any conclusion.

### Mental Health

Particular aspects of THR in mental health patients include a high prevalence of smoking and use of ENDS,^75,76,77^ preliminary evidence that ENDS are highly effective for smoking cessation in this group,^76^ and that patients with mental health issues may be more prone to addiction^78,79^ and struggle to quit nicotine in the longer term.^75^ Furthermore, nicotine itself may have an impact on symptoms and progression of the mental health condition.^80^

Seven studies investigating mental health outcomes were identified in this review, but there were others reporting on mental health disease as a predictor of ENDS use.^81,82^ The use of longitudinal or interventional study designs is even more crucial for mental health than for other health outcome scenarios due to the bi-directional link between mental health disorders and use of ENDS.

Of only two longitudinal studies on ENDS and mental health outcomes, one showed no deterioration in depressive symptoms and the other showed some deterioration, so no conclusion can be reached. Further studies are urgently required that are interventional in design and to investigate other health outcomes of switching from cigarettes to ENDS in this high-user patient group.

### Informing policy

The findings of this review show a failure of study designs to be able to rigorously establish causation with regards to health outcomes from the use of ENDS products.

The European Commission recently published its preliminary opinion on e-cigarettes stating that the weight of evidence is strong for risks of long-term systemic effects on the cardiovascular system, carcinogenicity of the respiratory tract and poisoning and injuries due to burns and explosion; moderate for local irritative damage to the respiratory tract and that other long-term adverse health effects, such as pulmonary disease, central nervous system and reprotoxic effects, cannot be established due to lack of consistent data.^83^ The findings of our systematic review do not support these conclusions.

Several of the studies included in this review that did not meet the criteria of being ‘good’ on strength of evidence or quality have been influential in determining health policy. One such study^42^ used data from the Population Assessment of Tobacco Health (PATH)^84^ database, which has also been used for several other articles included in this review.^20,48^ The study found that current e-cigarette users were twice as likely as never users to have had a MI.^42^ However, a subsequent reanalysis revealed that the majority of the 38 current e-cigarette users had their MI many years, on average a decade, before they first started using e-cigarettes.^85^ Despite the article being retracted by the publishing journal,^43^ the study findings had already been widely disseminated^86^ and cited,^87^ prior to its retraction with potential lasting impacts on the perception of CVD health risks from use of e-cigarettes.

Another incorrect health scare informing policy from use of ENDS occurred in 2019 with the “EVALI” outbreak which was initially widely reported as an outbreak of lipoid pneumonia due to vaping of nicotine.^88^ It was soon recognised and reported as being due to vaping of cannabinoid (THC) oils obtained from the black market rather than vaping of nicotine, with the CDC in the US recommending that adults using nicotine-containing e-cigarette, or vaping, products as an alternative to cigarettes should not go back to smoking.^88^

Our review found that very few studies were sufficiently rigorous to form conclusions on health risks and were not rigorous enough to inform policy on tobacco harm reduction.

The general public’s perception of health risks from ENDS does not reflect the available evidence and has become more negative according to the findings from two large surveys,^89^ whose authors underscored the urgent need to accurately communicate the risks of e-cigarettes to the public, which should clearly differentiate the absolute from the relative (to smoking) harms of e-cigarettes.

### Strengths and Limitations

This is the first systematic review to report on health outcomes for ENDS and has implications for use by policy makers. For example, current European policies requires packaging for ENDS products to report the same information on toxicity and addictiveness as for cigarettes and tobacco products.^90^ The European Commission has a consultation currently open on the health impact from ENDS.^83^ The findings of this study should form part of the scientific basis for such policies.

One limitation is that the authors assessing the quality of the studies were not blinded to the authors of the included studies, however standard protocols for systematic reviews were followed, with blinded independent reviews for level of evidence and quality, and very high (94%) inter-rater agreement.

We sought to identify only those articles where health outcomes from use of ENDS were the main research question. As such, our search was limited to searching for search terms in the title and our findings represent a reflection of the key studies in this field. The key health outcomes under investigation were mortality, CVD, respiratory and cancer as these make up the major health concerns from ENDS. We also searched for general health outcomes to identify the breadth of health outcomes being reported. There may be other research studies where health outcome was a secondary research question or fell outside of our search terms which may not have been captured in our study.

Our definition of ENDS did not include heat-not-burn devices, however a search using the same search protocol did not find any articles on the impact on health outcomes from heat-not-burn products.

We were unable to study the differential impact from various types of ENDS products and different constituent compounds (e.g., in nicotine fluid). In addition, different types of ENDS have different levels of nicotine delivery and addictive properties, which are likely to change the harmful effects (from components other than nicotine) of any product due to type of use (e.g. magnitude, time, etc.).

## Conclusion

This review of studies published over the last 5 years suggests the majority of studies on the use of ENDS products reported on negative health impacts with few reporting on health outcomes from switching from cigarettes to e-cigarettes. The strength of evidence and quality of the published studies is generally poor, yet some of these studies have been used to inform policy and are likely to have influenced public perception of health risks from use of ENDS. Several factors suggest the possibility of publication bias away from neutral or negative findings of harm to health outcomes from ENDS use.

Our review has demonstrated that ENDS use is not causative for any harmful CVD outcomes, and to the contrary, may be beneficial for hypertensive patients. Switching from cigarettes to e-cigarettes resulted in reduced exacerbations of COPD, with no evidence of long-term respiratory harm or deterioration in lung function. There was a suggestion from one study of short-term reductions in respiratory function in asthmatics, but no increased risk of asthma in ENDS users has been shown. Other health outcomes such as mental health, cancer and mortality have not been adequately studied to form a consensus on the health impact from ENDS use. However, the findings of our review did not negate the consensus held by many that ENDS use is safer than the risks posed from smoking cigarettes.

Overall, our review found very few studies were sufficiently rigorous to form conclusions on health risks. The research on ENDS use is not yet adequate to provide quantitative estimates about health risks. Consequently, the current body of evidence is inadequate for informing policy around tobacco harm reduction.

## Supporting information

Prisma Checklist

## Data Availability

This is a review. So, we do not have any primary data. We are only reporting already published data.

## Author Contributions

CH, ES and RP designed the study, conducted data extraction, analysis and review. CH and ES wrote the manuscript; SS, RN, PT and RP reviewed and edited the manuscript.

## APPENDIX 1

### SEARCH TERMS

Search terms for ENDS included: Electronic cigarette; Electronic nicotine delivery system; E-cigarette; Vaping; Vapor; Reduced risk tobacco product; Non cigarette tobacco; Nicotine aerosol; E-cigarette aerosol.

Search terms for health outcomes included: Health outcome; Morbidity; Mortality; Cancer; Cardiovascular disease; Chronic obstruct pulmonary disease; COPD; CVD; Acute myocardial infarction; Stroke; Cardiovascular; Cerebrovascular; Health effects; Adverse; effects; Respiratory.

## APPENDIX 2

### Abbreviations

BMI: Body mass index
CAT: COPD Assessment Test
CAD: Coronary artery disease
COPD: Chronic obstructive pulmonary disease
CVD: Cardiovascular disease
DBP: Diastolic blood pressure
ENDS: Electronic nicotine delivery systems
EC: Electronic cigarette
HR: Heart rate
MA: Meta-analysis
MI: Myocardial infarction
SBP: Systolic blood pressure

